# Depression and the Prefrontal-Hippocampal Pathway - A Pilot Multimodal Neuroimaging Study in Transgender Women

**DOI:** 10.1101/2024.10.14.24315485

**Authors:** Setthanan Jarukasemkit, Karen M. Tam, Seksan Yoadsanit, Ty Easley, Hailey Modi, Lyn Stahl, Adun Kampaengtip, Thanissara Chansakul, Rena Janamnuaysook, Akarin Hiransuthikul, Chaipat Chunharas, Janine D. Bijsterbosch

**Author notes:** Co-correspondence: **Chaipat Chunharas, MD, PhD**, Department of Neurology, Faculty of Medicine, Chulalongkorn University, Bangkok, Thailand., Address: 1873 Ratchadamri Rd., Pathum Wan, Bangkok 10330, Thailand, **Janine D. Bijsterbosch, PhD**, Department of Radiology, Washington University in St. Louis, Missouri, United States, Address: 4525 Scott Avenue, St. Louis, MO 63110, USA. joint last authors (Chaipat Chunharas, Janine D. Bijsterbosch).

## Abstract

This pilot study explored associations between gamma-aminobutyric acid (GABA) spectroscopy and functional magnetic resonance imaging in transgender women with depression. Twenty participants completed mental health surveys and imaging between January and March 2024. Depression severity was measured by Patient Health Questionnaire-9 (PHQ-9) scores. Higher PHQ-9 scores were associated with lower GABA levels in the hippocampus and lower fractional amplitude of low-frequency fluctuations (fALFF) in the dorsolateral prefrontal cortex. These findings revealed interactions between neurotransmitter signaling and functional brain activity of the hippocampal-prefrontal circuit in depression. Future large-scale neuroimaging studies are needed.

## Introduction

The transgender population experiences higher rates of depression compared to the cisgender population, with estimates in Thailand reaching around 22.7% for the transgender community compared to 3.2% in the overall population.^1^ Despite this high prevalence, transgender individuals are not typically represented in neuroimaging research projects aimed at elucidating the brain basis of depression. More broadly, there is a problematic lack of diversity in neuroimaging research, which largely focuses on cisgender participants from Western, Educated, Industrialized, Rich, and Democratic (WEIRD) countries.^2^

In this pilot study, we focused on two key brain regions implicated in depression, namely the hippocampus and the dorsolateral prefrontal cortex (DLPFC). Self-referential processes such as rumination, a key symptom of depression, have been associated with hippocampal hyperactivity, and hippocampal atrophy is commonly observed in recurrent depression.^3,4^ Furthermore, repetitive transcranial magnetic stimulation (TMS) applied to the left DLPFC has shown significant effectiveness in treating depression. ^5^ As such, the hippocampal-prefrontal circuit is of key interest in the context of depression.

A hierarchical model proposed by Hasler and Northoff has suggested that non-invasive measurement of cerebral gamma-aminobutyric acid (GABA) concentrations may offer endophenotypic insights into inhibitory pathways relevant to depression.^6^ A recent multimodal neuroimaging study showed that hippocampal GABA was necessary to recruit DLPFC activation in a task context that required suppression of unwanted thoughts.^7^ Despite the importance of the prefrontal-hippocampal circuit in cognitive and emotional regulation, its specific relationship with depression measurements remains poorly understood. To address these challenges with limited transgender representation in neuroimaging depression research and to elucidate the role of the prefrontal-hippocampal circuit in depression, the goal of this study was to investigate the prefrontal-hippocampal pathway of depression in a non-WEIRD cohort of transgender women.

## Material and Method

### Patients and Data Collection

Twenty transgender women with mild-to-severe depression were recruited from the Tangerine Community Health Clinic, a specialized healthcare center for transgender people in Bangkok, Thailand.^8^ Recruitment and data collection took place between January and March 2024. Mild-to-severe depression was defined based on Patient Health Questionnaire (PHQ-9) scores greater than 5 and a neuroticism score on the Five-Factor Inventory (NEO-FFI) greater than 30.1 which was previously defined as one standard deviation above the population mean. ^9,10^ Exclusion criteria comprised any history of neurological or cognitive disorders and contraindications for magnetic resonance imaging (MRI) scanning. See Supplementary Table S1 for a detailed overview of inclusion and exclusion criteria. Of the twenty subjects recruited, half were currently participating in gender-affirming hormone therapy (GAHT).

This study was approved by the Institutional Review Board, Faculty of Medicine Ramathibodi Hospital, Mahidol University (COA. MURA2023/522). After obtaining informed consent, participants completed a series of mental health questionnaire assessments followed by a one-hour MRI session at the Advanced Imaging Diagnostic Center, Faculty of Medicine Ramathibodi Hospital, Bangkok, Thailand. For the mental health assessments used in this manuscript, participants performed Thai language versions of the PHQ-9 and NEO-FFI self-report questionnaires on paper prior to the MRI scanning session. ^9,10^

### Neuroimaging Acquisition and Preprocessing

The MRI session included T1-weighted and T2-weighted structural MRI, resting-state functional MRI (rs-fMRI), and GABA magnetic resonance spectroscopy (MRS) imaging, using a Philips Ingenia Elition 3.0T scanner (software version R5.7.1). We leveraged a state-of the-art imaging protocol informed by the Human Connectome Project Lifespan and Disease protocols, and the MRS utilized the MEGA-PRESS sequence with parameters based on standard guidelines.^11^ For more details, the neuroimaging acquisition parameters are listed in Supplementary Table S2.

Functional imaging data were preprocessed by FMRIB’s Software Library (FSL) software. Briefly, the preprocessing steps included motion correction, distortion correction, structural co-registration, spatial normalization to Montreal Neurological Institute space using nonlinear registration, smoothing with a 5 millimeter kernel, and noise correction with independent component analysis. The distortion correction of echo planar imaging (EPI) of rs-fMRI was performed utilizing a technique that leverages two different phase encoding directions.^12^ Specifically, FSL’s topup tool estimated distortion fields from opposite phase-encoded EPI images, and the estimated fields were applied for geometric correction. Please see the Supplementary Methods for further detail. Additional cleanup of rs-fMRI data was performed using independent component analysis noise removal by FSL AROMA.

For MRS images, signals obtained from the left DLPFC, and left hippocampus were processed with baseline initial peak subtraction with 90% Gaussian character for fitting the power spectrum. MEGA Subtract All (ON-OFF) Spectra was performed. Please see the Supplementary Methods for further detail.

### Data Analysis

The study utilized neuroimaging parameters focused on two regions of interest: the hippocampus and the DLPFC. For the functional analyses, the bilateral hippocampus was defined based on the Harvard/Oxford Subcortical atlas and the bilateral DLPFC was defined based on prior work that utilized a task fMRI localizer (Figure 1).^13^ We extracted the mean time series of the left and right hippocampus and DLPFC following preprocessing and estimated the fractional amplitude of low-frequency fluctuation (fALFF). This measure captures the normalized strength of spontaneous signal fluctuations in each brain region by calculating the ratio of the power in the low-frequency range (0.01-0.1 Hertz) to the power across the full frequency range.^14^ Subsequently, the fALFF estimates were averaged between left and right hippocampus and between left and right DLPFC before further analysis.

**Figure 1:**
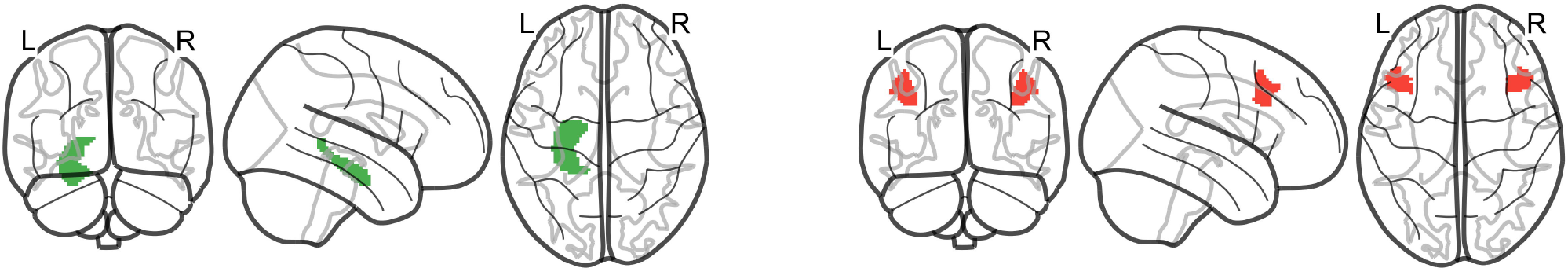
Regions of interest Regions of interest included the hippocampus (green) and DLPFC (red). L indicates the left hemisphere, and R indicates the right hemisphere.

For the spectroscopy data, GABA concentrations were normalized to creatinine concentrations (GABA/Cr) using creatinine as a stable reference metabolite. Notably, hippocampal spectroscopy can be challenging due to the small size of the hippocampus, which can lead to low spectral resolution and a low signal-to-noise ratio.^15^ To address this concern, we followed prior advice on the minimum voxel volume while also preserving spatial specificity for the regions of interest.^11^ Furthermore, we controlled for motion-related noise by including the framewise displacement parameter from rs-fMRI as a confound in subsequent analyses.

Following the extraction of neuroimaging parameters, multiple univariate linear regressions were performed using the ordinary least squares method. In these analyses, multimodal neuroimaging measures served as the independent variables, PHQ-9 scores were included as the dependent variables, and framewise displacement was included as a control variable. Mediation analyses were also conducted to investigate whether DLPFC fALFF mediated the relationship between hippocampal GABA and depression severity as measured with PHQ-9 scores. Additionally, an unpaired t-test was performed to investigate differences in neuroimaging measures and PHQ-9 scores between the GAHT and non-GAHT groups using permutation testing. Equations for all regression and mediation models can be found in the Supplementary Material.

## Results

The study consisted of 20 transgender women with mean age (standard deviation) of 30.1 (± 6.5) years. The mean neuroticism score (NEO-FFI) was 45.55 (± 7.5), and the mean PHQ-9 score was 14.6 (± 5.73), indicating the mental health status of the sample. Nearly half of the participants (45%) were undergoing gender-affirming hormone therapy at the time of the study. The linear regression models revealed a significant negative association between hippocampal GABA and PHQ-9 scores (β = -0.48, p = 0.039; Figure 2A), such that higher depression severity was associated with decreased hippocampal GABA. Furthermore, our findings revealed a significant negative association between DLPFC fALFF and PHQ-9 scores (β = -0.54, p = 0.015; Figure 2B), such that higher depression severity was associated with decreased fALFF in the DLPFC.

**Figure 2:**
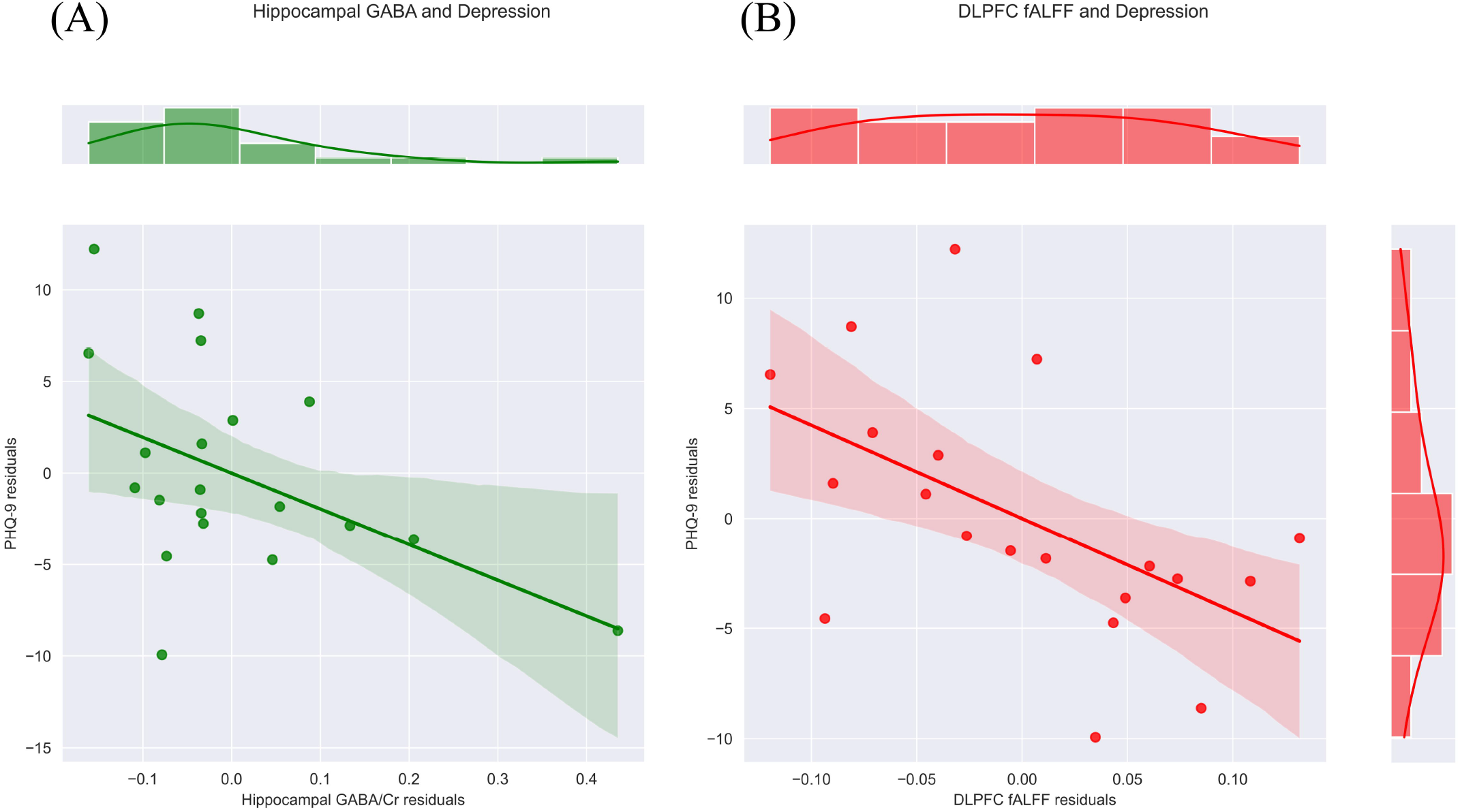
Associations between neuroimaging measures and depression severity Scatter plots illustrate the relationship of patient health questionnaire (PHQ-9) scores with gamma-aminobutyric acid (GABA) in the hippocampal (A) and fractional amplitude of low frequency fluctuation (fALFF) in the dorsolateral prefrontal cortex (DLPFC) (B) after controlling for the confounder. 95% Confidence intervals and marginal histograms are also displayed. GABA/Cr indicates GABA concentrations normalized to the creatinine reference.

No significant associations were observed between PHQ-9 scores and either hippocampal fALFF or DLPFC GABA.

Figure 3 illustrates the results from the mediation analysis model. We observed a trend-level mediating effect of DLPFC fALFF on the relationship between hippocampal GABA and PHQ-9 scores. Specifically, the results showed a significant relationship between hippocampal GABA and DLPFC fALFF (β = 0.48, p = 0.037) with a trend-level mediation effect (β = -0.20, p = 0.060).

**Figure 3:**
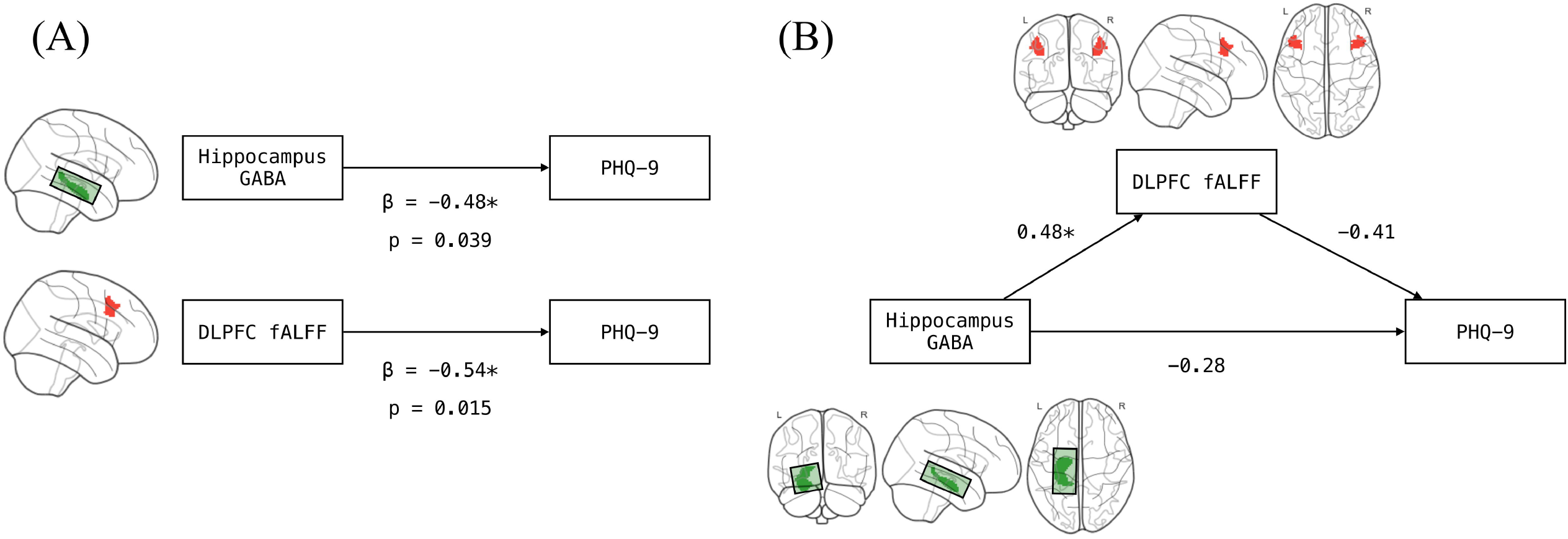
Path diagram of mediation analysis The right diagram displays separate linear models, while the left diagram illustrates the mediation analysis model. A) Direct associations between gamma-aminobutyric acid (GABA) in the hippocampus and patient health questionnaire (PHQ-9) scores, and between fractional amplitude of low frequency fluctuations (fALFF) in the dorsolateral prefrontal cortex (DLPFC) and PHQ-9 were estimated. B) The mediation analysis revealed a trend-level mediation effect of DLPFC fALFF on the relationship between hippocampal GABA and PHQ-9. The arrows indicate regression coefficients, and asterisks denote significance association (p < 0.05). L indicates the left hemisphere, and R indicates the right hemisphere.

We did not observe a significant difference between GAHT groups on PHQ-9 scores or on any of the neuroimaging measurements. Although prior work has shown a decrease in mental health symptomatology following GAHT, the lack of group differences in our study is explained by our inclusion criteria which required the presence of depressive symptoms to enable the investigation of the neural correlations of depression.^16^

## Discussion

This pilot study aimed to investigate the multimodal prefrontal-hippocampal circuit in relation to depression in a transgender cohort. Our findings revealed negative associations between depression severity and hippocampal GABA, and negative associations between depression severity and the amplitude of functional fluctuations in the DLPFC. These findings on the fronto-hippocampal relationship in depression are consistent with previous research in cisgender subjects, where higher hippocampal GABA was shown to recruit the DLPFC in an unwanted thought suppression task, indicating that DLPFC top-down signaling may depend on hippocampal GABA.^7^ Moreover, lower hippocampal GABA may be indicative of impaired inhibitory signaling, which could contribute to hippocampal hyperactivity.^3,17^

Hypoactivity of the DLPFC has previously been observed in major depressive disorder patients and in peripartum and post-stroke depression.^18,19,20^ Given the DLPFC’s pivotal role in cognitive control and emotion regulation, prior work has suggested that left DLPFC fALFF could serve as a biomarker to predict treatment responses to escitalopram and TMS intervention.^21,22^ Although GABA depletion is commonly observed in depression, GABA also plays critical roles in regulating neurochemical processes, including the hypothalamus-pituitary-adrenal axis and neurosteroid homeostasis.^23^ Disruption of GABA signaling in postpartum depression and premenstrual dysphoria may suggest abnormal neurochemical responses to hormonal fluctuations, particularly those driven by estradiol.^24,25^ As such, this link between estradiol and GABA may be of interest for future research on interactions between GAHT and depression in the transgender community.

Additionally, our study supports the Hasler & Northoff depression model, which links neurotransmitter imbalances, particularly in GABAergic signaling, to functional dysconnectivity.^6^ However, further research is required to explore all potential pathways within this model, including other circuitry like limbic and salience networks and other relevant neurotransmitters such as serotonin and dopamine. The findings of this pilot study provide preliminary validation for the potential use of GABA MRS in tracking symptom severity in this underrepresented population.

Although we were underpowered to detect differences linked to GAHT, GAHT has been shown to reduce the risk of depression in the transgender population and has previously been associated with lower odds of both depression and suicidality.^16^ These prior GAHT findings underscore the potential mediating role of hormone therapy in mitigating depression risk in the transgender population. Large-scale neuroimaging studies in transgender cohorts will be crucial to further understand the relationship between GAHT (and sex hormones more broadly) and neural circuitry dysfunction in depression.

### Limitation

The pilot study was limited in statistical power due to the relatively small sample size. As a result, we performed focused analyses on two brain regions (the hippocampus and DLPFC). Future large-scale multimodal neuroimaging studies that are sufficiently powered to interrogate a larger number of relationships in a broader neural circuitry are essential to address the complexities inherent in hierarchical models of depression.

## Conclusion

We investigated the prefrontal-hippocampal circuit in relation to depression in a transgender cohort using multimodal neuroimaging data. Our results revealed that higher depression severity was associated with reduced hippocampal GABA and with reduced DLPFC fALFF. Larger scale neuroimaging studies are needed to further study the brain basis of depression in the transgender community and the potential impact of GAHT on depression-related neural pathways.

## Data Availability

All data produced in the present study are available upon reasonable request to the authors.

## Author’s Contributions

The authors made the following contributions to this manuscript:

Conceptualization: SJ, TE, RJ, AH, CC, JDB;

Data curation: SJ, AK, TC;

Funding acquisition: SJ, TC;

Investigation: SJ, KMT, SY, TC;

Project administration: SJ;

Methodology: SJ, CC, JDB;

Validation: CC, JDB;

Software: CC, JDB;

Writing – original draft: SJ, CC, JDB;

Writing – review and editing: SJ, TE, HM, LS, RJ, AH, CC, JDB.

## Acknowledgements

We would like to express our gratitude to the Center for High Performance Computing and the Research Computing and Informatics Facility at Washington University in St. Louis for providing computational resources and technical support. We are also grateful to the Prince Mahidol Award Youth Program, Prince Mahidol Award Foundation under the Royal Patronage, Thailand for the financial support that enabled Dr. Jarukasemkit to conduct research at Washington University in St. Louis.

