## Supplementary Table S1 for "Depression and the Prefrontal-Hippocampal Pathway - A Pilot Multimodal Neuroimaging Study in Transgender Women"

Inclusion and Exclusion Criteria

All recruited participants were screened by Thai versions of Structured Clinical Interview for DSM-IV (SCID-IV), then filled self-administered two Thai questionnaires; Five-Factor Personality Inventory (NEO-FFI), and Patient Health Questionnaire-9 (PHQ-9). The inclusion criteria are in line with prior Human Connectome research.^1,2^

| **Inclusion Criteria** | **Exclusion Criteria** |
| --- | --- |
| Participants who exhibited sufficiently mild-to-severe depression defined by PHQ-9 > 5.^3,4^ Additionally, participant also exhibited depression personality trait defined by high neuroticism score, NEO-FFI Neuroticism > 30.1 (previously defined as one standard deviation above the population mean).^5,6^ | MRI contraindication (impairing claustrophobia, aneurysm clips, shunts, non-removable body piercings, non-removable cochlear or ear implants, permanent dentures or dental implants, joint replacements or prosthesis, pacemakers, defibrillators, or other implanted metal devices) |
| Transgender women who are Thai ancestry with age 18-60. | History of exclusionary neurological or cognitive disorder(s)/event(s) (amyotrophic lateral sclerosis, brain aneurysm, brain injury, brain tumors, cerebral palsy, Chiari malformation, dementia, encephalopathy, multiple sclerosis, Parkinson’s disease, recurrent epilepsy or seizure, stroke, or transient ischemic attacks) |

Note: Magnetic resonance imaging (MRI); NEO Five-Factor Inventory (NEO-FFI); Patient Health Questionnaire-9 (PHQ-9)
