## Supplementary Table S2 for "Depression and the Prefrontal-Hippocampal Pathway - A Pilot Multimodal Neuroimaging Study in Transgender Women"

Neuroimaging Acquisition Parameters

All modalities were scanned by Philips Ingenia Elition 3.0T scanner (software version R5.7.1) at the Advanced Imaging Diagnostic Center, Faculty of Medicine Ramathibodi Hospital, Bangkok, Thailand.

Details of acquisition parameters:

|  | **T1w** | **T2w** | **EPI** | **MEGA PRESS** |
| --- | --- | --- | --- | --- |
| TR | 9 ms | 2.5 s | 0.8 s | 1.6 s |
| TE | 4.2 ms | 330 ms | 37 ms | 68 ms |
| Flipped angle | 8 degrees | 90 degrees | 52 degrees | - |
| Voxel size | Isometric voxel  0.8x0.8x0.8 mm^3^ | Isometric voxel  0.8x0.8x0.8 mm^3^ | Isometric voxel  2.5x2.5x2.5 mm^3^ | Single voxel 4x2x3 cm^3^ for DLPFC and 4x2x2 cm^3^ for hippocampus |
| Slice thickness | 0.8 mm | 0.8 mm | 2.5 mm | - |
| Field of view | 256x256 mm^2^ | 256x256 mm^2^ | 208x208 mm^2^ | - |
| Fat suppression | ✓ | ✓ | - | - |
| Water excitation | - | - | - | ✓ |
| Bandwidth | 217 Hz/Px | 491 Hz/Px | 1813 Hz/Px | - |

Note: Echo planar imaging (EPI); Echo time (TE); Meshcher-Garwood point resolved spectroscopy (MEGA PRESS); Repetition time (TR), T1-weighted image (T1w); T2-weighted image (T2w); milliseconds (ms); millimeters (mm); seconds (s); centimeters (cm); Hertz per Pixel (Hz/Px).
