## Supplementary Methods for "Depression and the Prefrontal-Hippocampal Pathway - A Pilot Multimodal Neuroimaging Study in Transgender Women"

*Functional Magnetic Resonance Imaging (fMRI) Preprocessing*

We applied tools from the FMRIB Software Library (FSL)^1^ for all steps, including motion correction, distortion correction, structural co-registration, spatial normalization to Montreal neurological institute (MNI) space using nonlinear registration, smoothing, and independent component analysis (ICA).

Four subjects exhibited filler artifacts at the forehead; therefore, brain extraction was initially performed using BET over the time series. Motion correction was conducted using the FSL tool MCFLIRT^2^, and framewise displacement parameters were extracted using **fsl_motion_outliers** for use as confounders in the analysis.

Distortion correction was achieved through two echo planar imaging (EPI) sequences with opposite phase encoding directions^3^ using the **topup** and **applytopup** commands. As the data were acquired on a Philips scanner, effective echo spacing and total readout time were calculated as follows:

- Effective echo spacing = (((1000 * WFS)/(434.215 * (ETL + 1)))/acceleration)
- Total readout time = effective echo spacing * (ReconMatrixPE - 1)

WFS: water-fat shift (per pixel)
ETL: echo train length

Structural co-registration, spatial normalization, and smoothing were performed in the FSL tool FEAT. The EPI sequences for both phase encoding directions were analyzed separately, registered to the T1-weighted brain extracted image and standard space, MNI152_T1_2mm_brain, using a nonlinear method with a 10 mm warp resolution. Spatial smoothing was performed by full width half maximum 5 mm kernel, without temporal filtering.

Finally, ICA was performed using **fsl_melodic** in the FEAT directories. ICA-AROMA^4^ was then applied to identify noise components, and **fsl_regfilt** was used to remove noise components which resulted in the cleaned data.

*Functional Magnetic Resonance Imaging (fMRI) Postprocessing*

By applying masks, Harvard/Oxford Subcortical atlas^5^ for hippocampus, and prior mask for dorsolateral prefrontal cortex in MNI space^6^. Time series data were averaged inside each mask and fractional amplitude of low-frequency fluctuation^7^ were calculated for regions of interest.

*Magnetic Resonance Spectroscopy (MRS) Postprocessing*

Baseline initial peak subtraction with 90% Gaussian character for fitting the power spectrum and MEGA Subtract All (ON-OFF) Spectra were performed by Philips software. Hippocampal peak spectra showed gamma-Aminobutyric acid (GABA), Choline (Cho), Creatine (Cr), Phosphocreatine (Cr2), Glutamate + Glutamine (Glx), and N-Acetylaspartate (NAA).
