## Supplementary Materials for "Depression and the Prefrontal-Hippocampal Pathway - A Pilot Multimodal Neuroimaging Study in Transgender Women"

Supplementary Material

Linear Regression Models

Y = β_0_ + β_1_* X_1_ + β_5_*C + ε

Y = β_0_ + β_2_* X_2_ + β_6_*C + ε

Y = β_0_ + β_3_* X_3_ + β_7_*C + ε

Y = β_0_ + β_4_* X_4_ + β_8_*C + ε

Y is a dependent variable, which is Patient Health Questionnaire-9 (PHQ-9).

X_1_, X_2_, X_3_, X_4_ are independent variables, which are hippocampal ratio of gamma-Aminobutyric acid to Creatine (GABA/Cr), dorsolateral prefrontal cortex (DLPFC) GABA/Cr, hippocampal fractional amplitude of low-frequency fluctuations (fALFF), and DLPFC fALFF, respectively.

C is a control variable, which is framewise displacement.

β_1_, β_2,_ β_3_, β_4,_ β_5_, β_6,_ β_7_, β_8_ are linear regression coefficients.

β_0_ is a constant.

ε is residuals.

Mediation Analysis Equation

Y = β_1_*X + β_5_*C + c_1_ + ε_1_

M = β_2_*X + β_6_*C + c_2_ + ε_2_

Y = β_3_*X + β_4_*M + β_7_*C + c_2_ + ε_3_

Y is a dependent variable, which is PHQ-9.

X is an independent variable, which is GABA/Cr.

M is a mediator variable, which is DLPFC fALFF.

C is a control variable, which is framewise displacement.

β_1_, β_2_, β_3_ are linear regression coefficients, and β_3_ is direct effect (indirect effect equals to β_2_ * β_4_).

c_1_, c_2_, c_3_ are constants.

ε_1_, ε_2_, ε_3_ are residuals.
